# Durability of immunity and clinical protection in nursing home residents following bivalent SARS-CoV-2 vaccination

**DOI:** 10.1101/2023.04.25.23289050

**Authors:** Stefan Gravenstein, Frank DeVone, Oladayo A. Oyebanji, Yasin Abul, Yi Cao, Philip A. Chan, Christopher W. Halladay, Kevin W. McConeghy, Clare Nugent, Jürgen Bosch, Christopher L. King, Brigid M. Wilson, Alejandro B. Balazs, Elizabeth M. White, David H. Canaday

## Abstract

**Background:** Vaccines have substantially mitigated the disproportional impact of SARS-CoV-2 on the high morbidity and mortality experienced by nursing home residents. However, variation in vaccine efficacy, immune senescence and waning immunity all undermine vaccine effectiveness over time. The introduction of the bivalent vaccine in September 2022 aimed to counter this increasing susceptibility and consequences of breakthrough infection, however data on the durability and protection of the vaccine are limited. We evaluated the durability of immunity and protection after the first bivalent vaccination to SARS-CoV-2 in nursing home residents.

**Methods:** For the immunologic evaluation, community nursing home volunteers agreed to serial blood sampling before, at two weeks, three and six months after each vaccination for antibodies to spike protein and pseudovirus neutralization activity over time. Concurrent clinical outcomes were evaluated by reviewing electronic health record data from residents living in Veterans Administration managed nursing home units. Residents without recent infection but prior vaccination to SARS-CoV-2 were followed over time beginning with administration of the newly available bivalent vaccine using a target trial emulation (TTE) approach; TTE compared time to breakthrough infection, hospitalization and death between those who did and did not receive the bivalent vaccine.

**Results:** We evaluated antibodies in 650 nursing home residents; 452 had data available following a first monovalent booster, 257 following a second monovalent booster and 321 following a bivalent vaccine. We found a rise in BA.5 neutralization activity from the first and second monovalent boosters through the bivalent vaccination regardless of prior SARS-CoV-2 history. Titers declined at three and six months after the bivalent vaccination but generally exceeded those at three months compared to either prior boost. BA.5 neutralization titers six months after the bivalent vaccination were diminished but had detectable levels in 80% of infection-naive and 100% of prior infected individuals. TTE evaluated 5903 unique subjects, of whom 2235 received the bivalent boost. TTE demonstrated 39% or greater reduction in risk of infection, hospitalization or death at four months following the bivalent boost.

**Conclusion:** Immunologic results mirrored those of the TTE and suggest bivalent vaccination added substantial protection for up to six months after bivalent vaccination with notable exceptions. However, the level of protection declined over this period, and by six months may open a window of added vulnerability to infection before the next updated vaccine becomes available. We strongly agree with the CDC recommendation that those who have not received a bivalent vaccination receive that now and these results support a second bivalent booster for those at greatest risk which includes many nursing home residents.

## Introduction

SARS-CoV-2 continues to cause disproportionate morbidity and mortality in older adults (1). Although vaccines effectively reduce this burden, immunity wanes in the months following vaccination (2, 3) and the virus continues to evolve to escape population immunity (4, 5). As newer versions of vaccines are adapted to recent SARS-CoV-2 variants (i.e., bivalent vaccine)(6), we must continue to determine how effective these vaccines are over time and against emerging variants and especially in vulnerable populations.

One vulnerable population to study both immunologic and clinical measures of protection include nursing home residents. Nursing homes have suffered some of the most significant consequences of SARS-CoV-2 infection as measured by mortality in its residents and workers (7), due to the close proximity of living arrangements, significant care needs that increase person-to-person contact amplifying the opportunity for transmission, and house a population with multiple morbidities, frailty and senescent immune systems (8). It is therefore critical to evaluate both immunologic correlates of immunity and clinical outcomes such as infection, hospitalization and death following vaccination in this population.

Our cross-institution collaboration provided a unique opportunity to evaluate these outcomes concurrently in two separate populations, immunologic responses to vaccine in community nursing home (CNH) residents, and clinical outcomes in nursing home residents of care managed by the Veterans Administration, i.e., in community living centers (CLCs), the CNH equivalent for Veterans. Here, we report on durability of protection following the first bivalent SARS-CoV-2 vaccine first offered September 2022, measured by immunity in CNH, and clinical outcomes for CLC residents followed for infection, hospitalization, and mortality.

## Methods

### Ethical Approval

The immunology study requiring individuals consenting was approved by the Western Copernicus Group national institutional review board. All participants or their legally authorized representatives provided informed consent. The Providence VA Healthcare System IRB approved the protocol ‘COVID-19 in VA Community Living Centers’ and waived informed consent for the component of this work using a target trial emulation (TTE) approach.

### Immunology

#### Participants

Residents were sampled from community nursing homes in Ohio and Rhode Island and two state Veterans Homes. All sites administered the BNT162b2 or mRNA-1273 COVID-19 vaccines. The vast majority of subjects however received BNT162b2 mRNA, the vaccine predominantly offered to nursing home residents. Subjects received the primary series, first, second and then bivalent boosters shortly after they were authorized by FDA and recommended by CDC’s Advisory Committee on Immunization Practices (ACIP).

Participants were deemed to have a “*prior infection*” if they had a known history of SARS-CoV-2 infection confirmed by PCR or antigen test, and/or detectable antibody levels to SARS-CoV-2 spike, receptor binding domain (RBD), and Nucleocapsid (N protein) from serum collected prior to their first dose in the initial study similar to our prior studies. (9, 10). Otherwise, participants were classified as “*infection-naive*.” Throughout the course of the longitudinal study including the multiple vaccinations, if a subject was PCR/antigen positive and/or had a rise outside of laboratory variance of anti-spike, RBD, N-protein, and neutralizing assay results not accounted for by vaccination history, they were deemed to have had an infection whether clinically detected or not. Their subsequent data points were then re-classified from that time point onward.

Serum samples were obtained at multiple time points after each vaccination including at two weeks, and then three- and six-months.

#### Anti-spike and anti-N assay

Immune response to the vaccine was assessed using a bead-multiplex immunoassay using Wuhan strain and BA.5 (10). Anti-spike IgG generated a result of BAU/ml based on the Frederick National Laboratory standard which was calibrated to the WHO 20/136 standard for Wuhan. Anti-spike BA.5 uses arbitrary units (AU/ml). Stabilized full-length spike protein (aa 16-1230, with furin site mutated) of each strain was used. For the full-length Wuhan, N-protein cutoffs for positivity and N changes were used to assess prior infection or breakthrough as done previously (10, 11).

#### SARS-CoV-2 pseudovirus neutralization assay

To determine the neutralizing activity of vaccine recipients’ sera against coronaviruses, we produced lentiviral particles pseudotyped with spike protein based on the Wuhan and Omicron BA.5 strain as previously described (10, 12). Briefly, neutralization assays were performed using a Fluent 780 liquid handler (Tecan) in 384-well plates (Grenier). Three-fold serial dilutions of serum ranging from 1:12 to 1:8,748 were performed and added to 50–250 infectious units of pseudovirus for one hour. pNT50 values were calculated by taking the inverse of the 50% inhibitory concentration value for all samples with a pseudovirus neutralization value of 80% or higher at the highest concentration of serum. The lower limit of detection (LLD) of this assay is 1:12 dilution.

### Statistical Methods for immunology studies

Subjects with Wuhan or BA.5 Spike antibody or neutralizing titers for at least one sample between pre-1st monovalent booster and six months after the bivalent vaccination were included in the immunology analysis. Samples that coincided with a breakthrough infection were excluded. Among the remaining subjects, demographics were summarized overall and separately for each vaccine dose.

For each sample time, the geometric mean titer (GMT) was calculated by assay and strain and separately among infection-naive and prior infection subjects at the given time. Given the repeated sampling of subjects and missing data due to enrollment, disenrollment/discharge, breakthrough infections, or subject unavailability, mixed-effects models were used to compare titer levels while adjusting for correlated observations and unbalance across doses and follow-up times.

Using the dates of the draws relative to the last vaccine dose, random-intercept models were estimated by assay and strain predicting log-transformed titer response as a function of days since vaccine dose with a 2nd-degree polynomial function and considering subjects as a random effect. Different polynomial functions were allowed for each vaccine dose and the prior vs. naive status of the subject at the draw. To compare the post-boost titer levels of the three vaccinations (two boosters and bivalent vaccine) to each other and to the six-month post-bivalent levels, additional mixed-effects models were estimated. Random intercepts were estimated for each subject four times (post-1st monovalent, post-2nd monovalent, post-bivalent, and six months post-bivalent) were compared for each assay, strain, and naive/prior status. Models and contrasts were estimated using the nlme and emmeans packages in R4.2.2 (13, 14).

### Clinical TTE Methods

We deployed a Target Trial Emulation (TTE) approach, similar to what we reported for clinical outcomes following monovalent boosters (15, 16), to assess additional clinical effectiveness following the introduction of a bivalent SARS-CoV-2 vaccine in September 2022. On each day from September 18 on we evaluated if Veterans living in a Veterans Administration-managed nursing home, i.e., Community Living Center or CLC, were eligible for inclusion. Subjects eligible for inclusion had to be living in a CLC facility for at least 100 days, with a gap of no more than ten days (i.e., residents could leave and come back, but not leave for >10 days), and have received the primary series. We excluded those who had a test-confirmed SARS-CoV-2 infection within the prior 90 days, a SARS-CoV-2 vaccine within the prior 134 days or in hospice care. Trial dates included those index dates for eligible subjects between September 18, 2022 and March 30, 2023, and follow-up was for up to March 31, 2023. Those in the exposed arm were indexed on their date of bivalent administration, those in the control arm had their index randomly selected from among eligible dates where at least one member of the exposed group received a bivalent vaccination. Follow-up was censored upon outcome, leaving the CLC (and not returning in <10 days), death (when death is not the outcome) or March 31, 2023, whichever comes first.

Clinical outcomes included a PCR test-confirmed SARS-CoV-2 infection, death within 30 days or hospitalization within 14 days of SARS-CoV-2 infection confirmation, or a combination of hospitalization or death in these intervals.

The initial analysis at the Veteran-Trial level uses a Kaplan-Meier estimator with cumulative incidence for each group for unadjusted hazard ratios. We calculated unadjusted and adjusted results to the Veteran Trial Time (day) level and emulate the equivalent of running a trial every single day until a censoring event or outcome event occurs per veteran-trial-day, again mimicking our previously reported results (16). We calculate joint inverse probability weights for both treatment and censoring, with our final results being the cumulative incidence at specific time points representing vaccine effectiveness at weeks 12 and 16. Model selection for weights was based on a priori experience with confounders (i.e., initial booster paper, subsequent vaccine work) and observed demographic and diagnosed data. Weights were truncated at their 95% upper quantile. Sampling with replacement by resident (i.e., bootstrapping) with 500 replications was used to generate 95% confidence intervals accounting for the estimated probability weights and crossover of treatment groups by resident. All TTE analyses were performed using R statistical software, version 4.2.2 (R Foundation for Statistical Computing). The main difference from our previously reported results was the inclusion of trial days without full follow up, as such this approach produces longer follow-up periods for those who have earlier index dates, and much shorter ones for those with later index dates.

## Results

### Neutralization and anti-spike titers after boosting

This immunology study is of a cohort of long-stay nursing home residents who received the COVID-19 mRNA vaccine primary series, two monovalent boosters, and now one bivalent vaccination. Table 1 describes the numbers and demographics of the subjects at each vaccine dose. Not all subjects were drawn or followed through all the time points. Follow-up blood sampling was performed two weeks after each dose for a peak response and then three and six months following each vaccine to determine how sustained the response was. We also received clinical information in the follow-up intervals to assess if they acquired SARS-CoV-2 infection detected clinically either through symptomatic testing, outbreak screening to identify asymptomatic persons or our serologic studies. If clinical or laboratory data supported a recent infection, we excluded their data until they received their next vaccine dose and converted their clinical category from naive to prior infected if they were not prior infected already. In our recent initial report on the bivalent vaccination, 77% of the nursing home cohort had evidence of prior infection (10). The current study also included a group of residents that appeared to have never been infected, allowing us to perform a subgroup analysis of naive and prior infected residents. They have distinctly different responses.

**Table 1.** Demographics for immunology studies.

| Subjects | All | Booster 1 | Booster 2 | Bivalent |
| --- | --- | --- | --- | --- |
| Number | 650 | 452 | 257 | 321 |
| Age years (Med IQR) | 76 (68,86) | 76 (69,85) | 76 (68,86) | 74 (67,85) |
| Age Range | 33-106 | 33-106 | 33-103 | 33-104 |
| Male | 335 (52%) | 254 (56%) | 115 (45%) | 155 (48%) |
| Female | 315 (48%) | 198 (44%) | 142 (55%) | 166 (52%) |
| Race: White | 515 (79%) | 354 (78%) | 206 (80%) | 253 (79%) |
| Black | 118 (18%) | 86 (19%) | 43 (17%) | 62 (19%) |
| Hispanic | 8 (1%) | 6 (1%) | 5 (2%) | 2 (1%) |
| Asian | 2 (0%) | 2 (0%) | 0 (0%) | 0 (0%) |

The primary focus of this study was on the six-month timepoint post-bivalent vaccination. We report neutralizing and anti-spike titers to both components of the bivalent vaccine - BA.5 and Wuhan. The neutralizing assays (Figure 1 and Table 2) demonstrate that with each vaccine dose from the first two monovalent and then the bivalent vaccination, both BA.5 and Wuhan neutralization titer rises in both naive and prior infection groups. Table 3 demonstrates the ratio of change from one vaccination to the next of the immune parameters with the neutralization assay changes having the highest and most significant elevations from dose to dose in the infection-naive and prior infected groups. At virtually all time points, the prior infected individuals have higher titers than the naive group (Figure 1). The levels decline after three and six months as expected at all time points noted in Figure 1 and statistically reported for six months post-bivalent vaccination in Table 2. The BA.5 neutralization level decreased by >90% (fold change = 0.07) in naive and >85% in prior infection by six months after the bivalent vaccination. However, 80% (9/12) of persons who are infection-naive still had detectable BA.5 neutralization activity while 100% (32/32) had detectable levels in those with prior infection.

**Figure 1.**
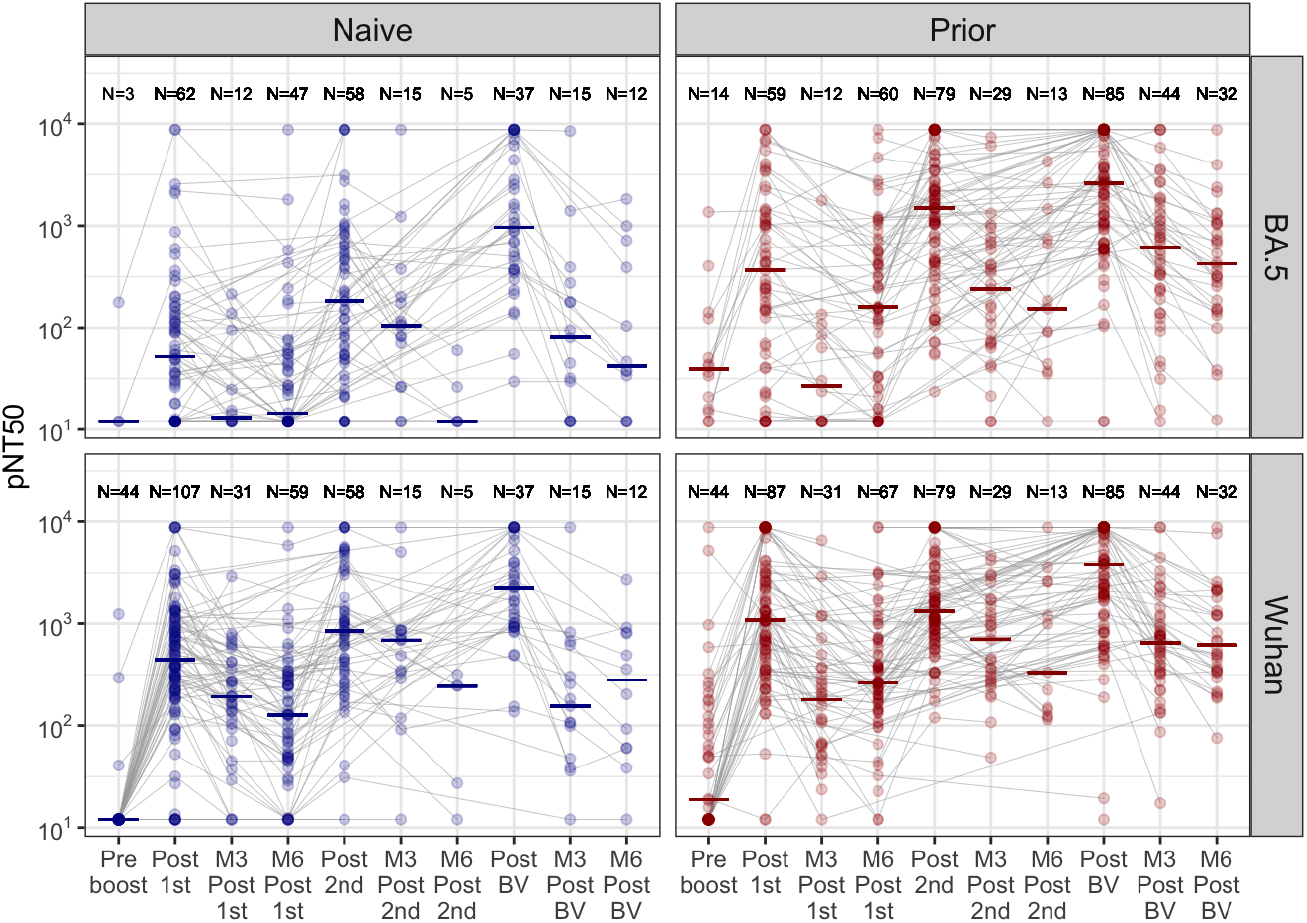
Neutralization assays after boosting. This figure illustrates pseudovirus neutralization assay results for Omicron BA.5 (top panels), and Wuhan strains (bottom panels) in nursing home residents before and after first and second monovalent booster and the bivalent booster. The blue panels are infection-naive and the red panels had prior infection. Not all subjects had all timepoints drawn. N is the subjects in that group. The solid line is the median response. pNT50 = Pseudovirus neutralization. X axis timepoints: Preboost is 0-14 days prior the the first monovalent booster, Postboost 1st, 2nd, BV are a median 17 days, M3 Post is median 105 days, and M6 Post is median 183 days.

**Table 2.** Titers of post boosts and 6 month post bivalent booster.

| Strain | Status | Post 1st<br>GMT (CI) | Post 2nd<br>GMT (CI) | Post BV<br>GMT (CI) | 6 months post<br>BV GMT (CI) | Adjusted Ratio:<br>6 months/<br>2 week Post BV | p value of<br>ratio |
| --- | --- | --- | --- | --- | --- | --- | --- |
| **Neutralization** |  |  |  |  |  |  |  |
| BA.5 | Naive | 68<br>(43, 109) | 195<br>(120, 318) | 1136<br>(671, 1921) | 88<br>(28, 278) | 0.066 | <0.001 |
| BA.5 | Prior | 299<br>(174, 514) | 1181<br>(840, 1660) | 2075<br>(1518, 2835) | 424<br>(257, 700) | 0.16 | <0.001 |
| Wuhan | Naive | 425<br>(313, 578) | 840<br>(565, 1247) | 2068<br>(1428, 2994) | 214<br>(77, 591) | 0.13 | <0.001 |
| Wuhan | Prior | 1073<br>(778, 1479) | 1487<br>(1174, 1883) | 2701<br>(2026, 3599) | 740<br>(498, 1100) | 0.25 | <0.001 |
| **Spike** |  |  |  |  |  |  |  |
| BA.5 | Naive | 2228<br>(1397, 3552) | 968<br>(679, 1379) | 1216<br>(943, 1567) | 417<br>(196, 888) | 0.26 | <0.001 |
| BA.5 | Prior | 3500<br>(2301, 5324) | 2093<br>(1683, 2602) | 1390<br>(1180, 1637) | 1250<br>(914, 1711) | 0.61 | 0.016 |
| Wuhan | Naive | 1954<br>(1430, 2672) | 1649<br>(1046, 2601) | 3267<br>(2375, 4493) | 412<br>(195, 869) | 0.087 | <0.001 |
| Wuhan | Prior | 6429<br>(5210, 7934) | 3744<br>(3070, 4565) | 3282<br>(2736, 3938) | 1107<br>(797, 1537) | 0.25 | <0.001 |

Geometric mean titer (GMT), Confidence interval (CI), Bivalent (BV), Month 6 (M6), Adjusted Ratio of Month 6 PostBV / Post BV calculation will be slightly different from the crude ratios of the presented GMTs because the mixed-effects model adjusts for correlated values within subject when > 1 sample is present from the same person. The GMT columns ignore repeated sampling and consider all values as independent of each other. BV = bivalent vaccination, GMT= geometric mean titer, Status means whether infected before vaccination, "Prior" means in individuals who had been infected with SARS-CoV-2 before vaccination, and "Naive" means individuals who have not been infected with SARS-CoV-2 before vaccination.

**Table 3.**
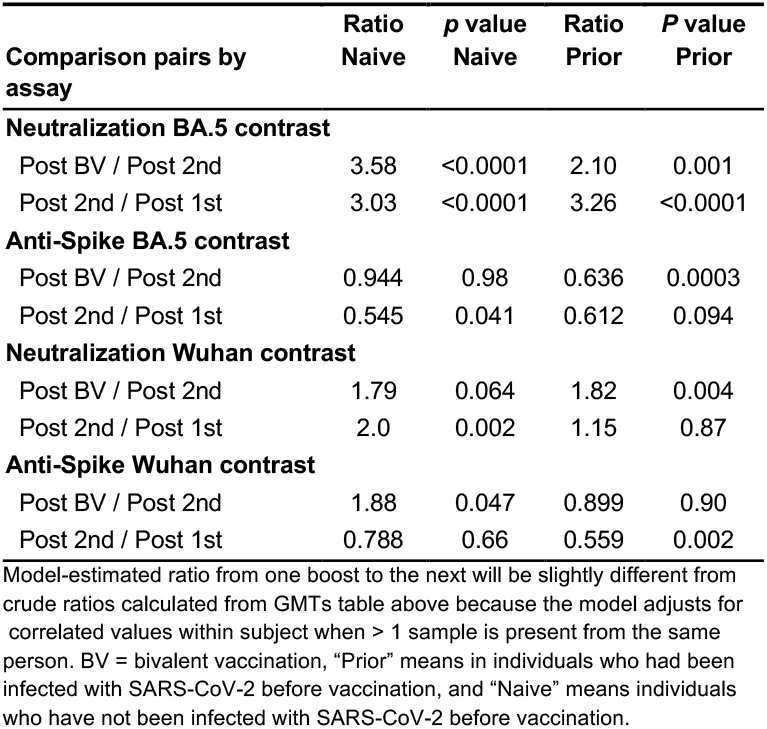
Model-estimated contrasts of ratio of change after each booster.

| Comparison pairs by<br>assay | Ratio<br>Naive | p value<br>Naive | Ratio<br>Prior | P value<br>Prior |
| --- | --- | --- | --- | --- |
| **Neutralization BA.5 contrast** |  |  |  |  |
| Post BV / Post 2nd | 3.58 | <0.0001 | 2.10 | 0.001 |
| Post 2nd / Post 1st | 3.03 | <0.0001 | 3.26 | <0.0001 |
| **Anti-Spike BA.5 contrast** |  |  |  |  |
| Post BV / Post 2nd | 0.944 | 0.98 | 0.636 | 0.0003 |
| Post 2nd / Post 1st | 0.545 | 0.041 | 0.612 | 0.094 |
| **Neutralization Wuhan contrast** |  |  |  |  |
| Post BV / Post 2nd | 1.79 | 0.064 | 1.82 | 0.004 |
| Post 2nd / Post 1st | 2.0 | 0.002 | 1.15 | 0.87 |
| **Anti-Spike Wuhan contrast** |  |  |  |  |
| Post BV / Post 2nd | 1.88 | 0.047 | 0.899 | 0.90 |
| Post 2nd / Post 1st | 0.788 | 0.66 | 0.559 | 0.002 |

Model-estimated ratio from one boost to the next will be slightly different from crude ratios calculated from GMTs table above because the model adjusts for correlated values within subject when > 1 sample is present from the same person. BV = bivalent vaccination, "Prior" means in individuals who had been infected with SARS-CoV-2 before vaccination, and "Naive" means individuals who have not been infected with SARS-CoV-2 before vaccination.

A focused examination of the decay of neutralization titers over six months in each post-vaccination category is shown (Figure 2). The decays after each vaccination follow a similar parallel pattern, with the prior infection group having higher titers at the start that stay higher than in infection-naive individuals throughout the six months after each vaccination. The decay curves in both groups from the three- to six-month timepoints after the bivalent vaccination have a relative flattening with the anti-BA.5 spike (Table 2 and Figure 3) with readily detectable levels both explain the maintenance of some degree of BA.5-specific neutralization through at least six months in most individuals and even those that are infection-naive.

**Figure 2.**
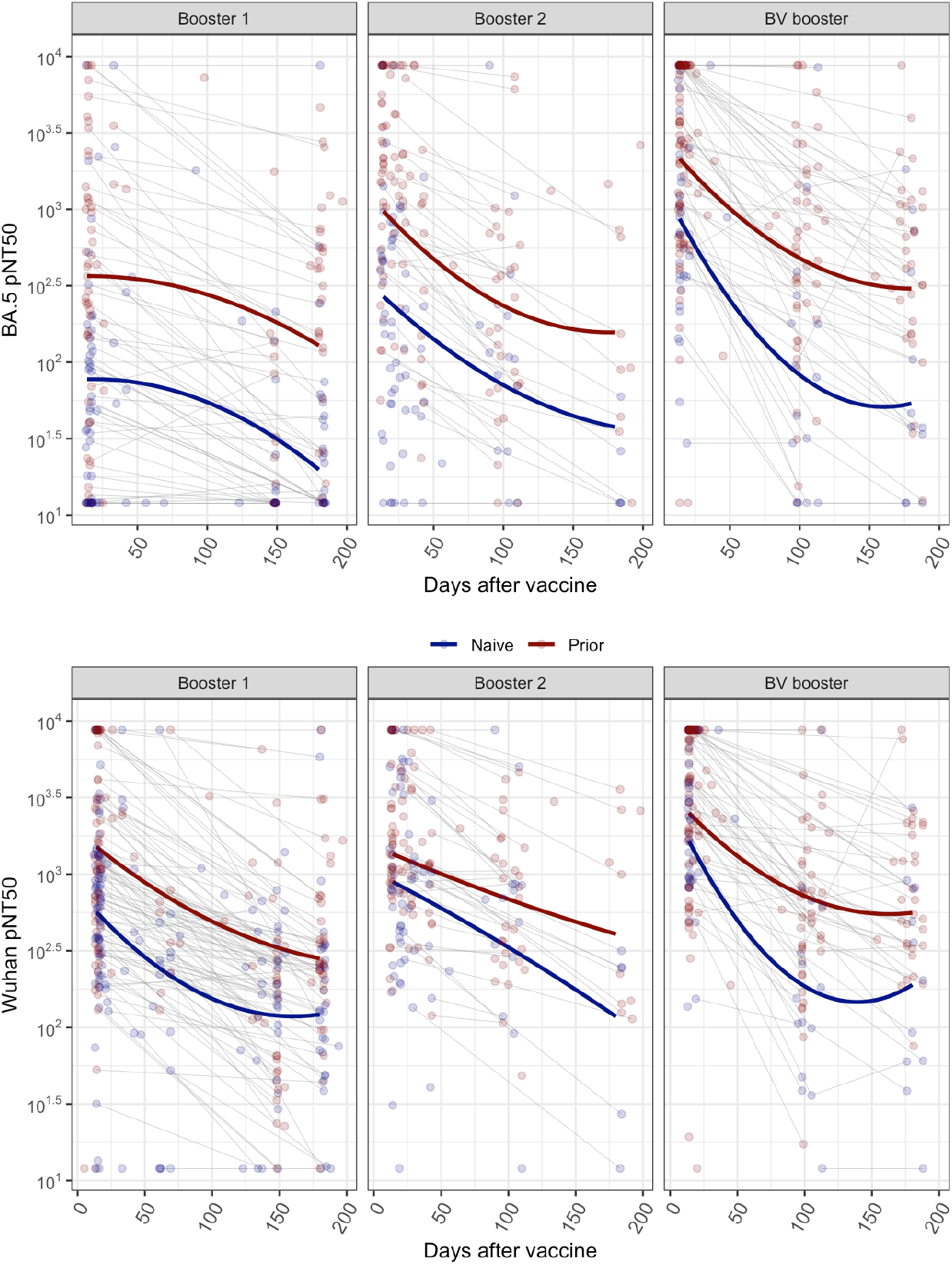
Decline of neutralization titers after boosting. pNT50 neutralization titer of BA.5 (top) and Wuhan (bottom) of days from each booster dose in infection naive (blue) and prior infected (red). The lines represent the estimated fixed effects from a mixed-effects model predicting log-transformed titer response as a function of days since vaccine dose with a 2nd degree polynomial function and random intercepts at the subject level.

**Figure 3.**
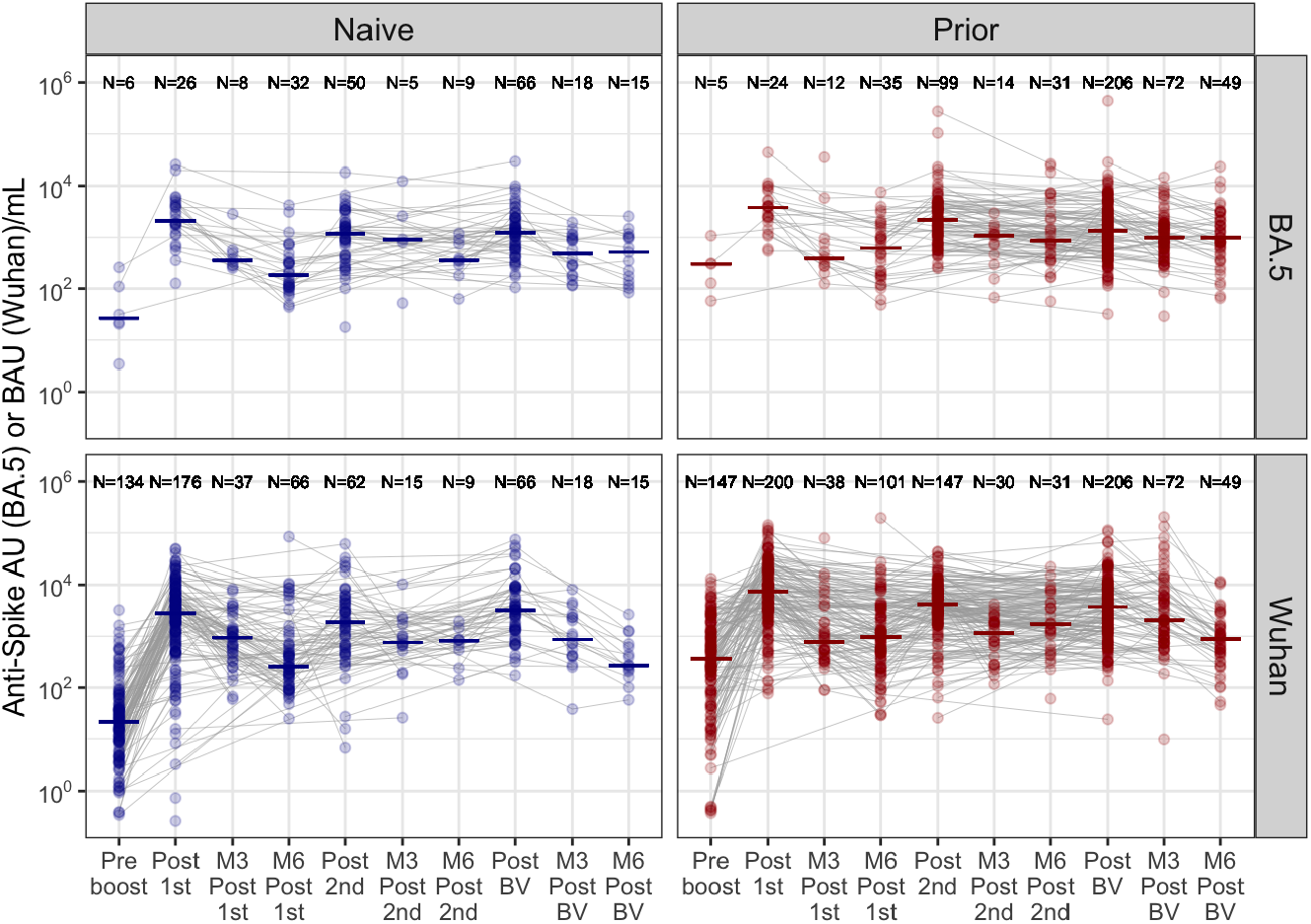
Anti-spike titers after boosting. This figure illustrates anti-spike for BA.5 (top) in AU/ml and Wuhan (bottom) in BAU/ml in nursing home residents before and after first and second monovalent booster and the bivalent booster. The blue panels are infection-naive and the red panels had prior infection. Not all subjects had all timepoints drawn. N is the subjects in that group. The solid line is the median response. X axis timepoints: Preboost is 0-14 days prior the the first monovalent booster, Postboost 1st, 2nd, BV are a median 17 days, M3 Post is median 105 days, and M6 Post is median 183 days.

The anti-spike binding assay titers reveal that all subjects regardless of their prior infection category have a readily detectable level of anti-BA.5 and Wuhan spike titers (Figure 3). The binding titers in contrast to neutralization titers reach a peak level of the first boost and do not continue to rise with subsequent boosts while the BA.5 and Wuhan neutralization titers do rise from boost to boost (Table 3). The neutralization assay is a more functional assay. Interestingly even in the presence of lesser anti-spike levels, the neutralization titer still rises with each vaccine dose. Also, the relative difference between the naive and prior infected group in anti-spike titers is blunted compared to the differences noted in neutralization titers.

### TTE Results

The full cohort of residents eligible between 9/18/22 to 3/18/23 includes 3,783 persons with 5,903 observations (Veteran-trials), across 127 CLCs. Table 4 shows baseline covariates, and that most Veterans had received at least one boost. Disproportionately more of those who received a bivalent vaccine also had two prior boosts, and influenza vaccination (Table 4). Reduced risk for infection (Figure 4 and 5), hospitalization and death is evident in the first months following bivalent vaccination (Table 5 and Figure 5). Available follow-up time from vaccination dictates how many individuals remain available for determination of efficacy estimates, and the last available data as of March 31, 2023 left less than 10% of our bivalent vaccinated sample with more than four months of follow-up. Nevertheless, bivalent vaccination appears to provide benefit from infection for at least three months (41% reduction; 95% CI 24.7-54.7%), and hospitalization (47.2%; 95% CI 22.3-67.6%) or the composite of hospitalization and death for somewhat longer (50.5%; 95% CI 26.1-68.6%) (Table 5 and Figure 5). Too few people died for the estimated 74.4% reduction in death at 16 weeks to reach statistical significance.

**Table 4.**
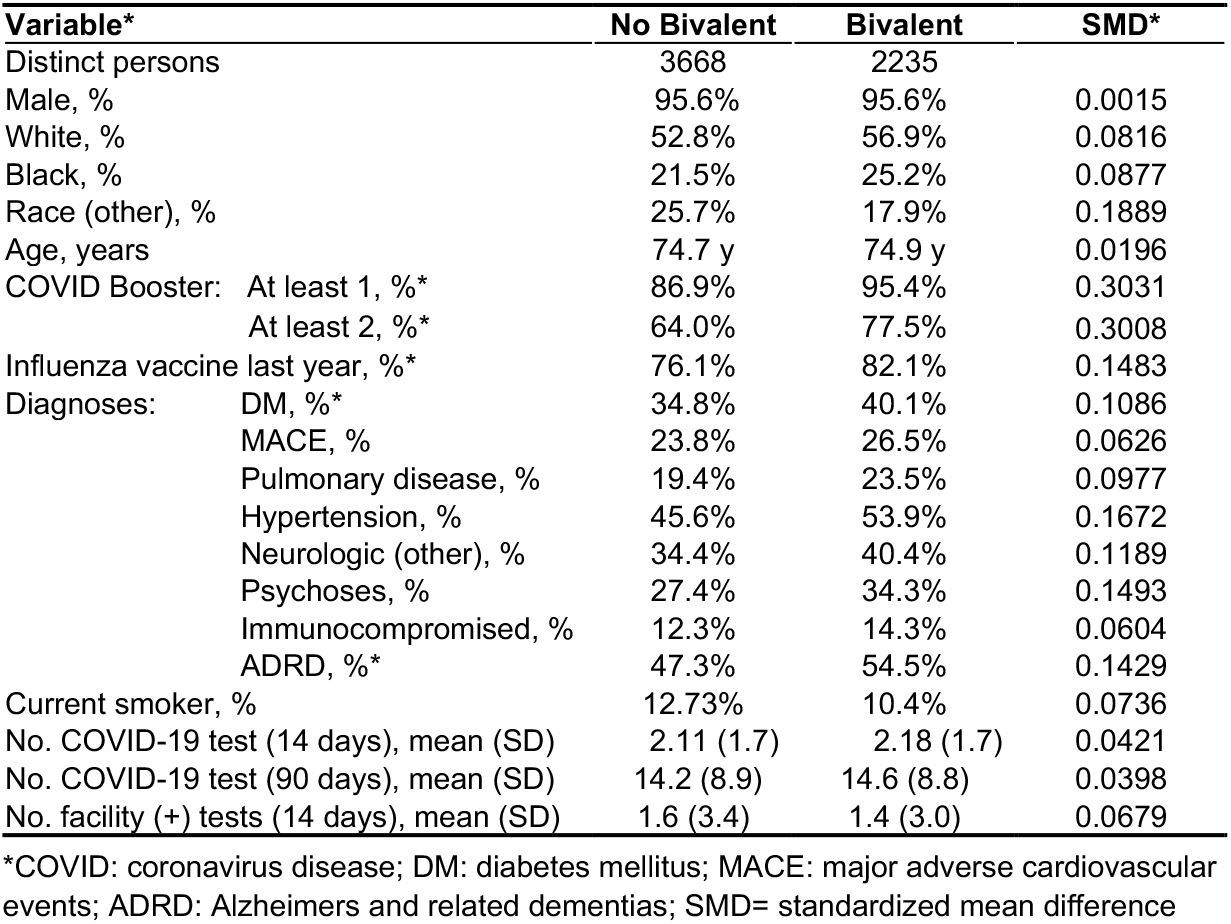
Selected baseline co-variates for target trial emulation population, and those with SMD >0.10.

| Variable* | No Bivalent | Bivalent | SMD* |
| --- | --- | --- | --- |
| Distinct persons | 3668 | 2235 |  |
| Male, % | 95.6% | 95.6% | 0.0015 |
| White, % | 52.8% | 56.9% | 0.0816 |
| Black, % | 21.5% | 25.2% | 0.0877 |
| Race (other), % | 25.7% | 17.9% | 0.1889 |
| Age, years | 74.7 y | 74.9 y | 0.0196 |
| COVID Booster: At least 1, %* | 86.9% | 95.4% | 0.3031 |
| At least 2, %* | 64.0% | 77.5% | 0.3008 |
| Influenza vaccine last year, %* | 76.1% | 82.1% | 0.1483 |
| Diagnoses: DM, %* | 34.8% | 40.1% | 0.1086 |
| MACE, % | 23.8% | 26.5% | 0.0626 |
| Pulmonary disease, % | 19.4% | 23.5% | 0.0977 |
| Hypertension, % | 45.6% | 53.9% | 0.1672 |
| Neurologic (other), % | 34.4% | 40.4% | 0.1189 |
| Psychoses, % | 27.4% | 34.3% | 0.1493 |
| Immunocompromised, % | 12.3% | 14.3% | 0.0604 |
| ADRD, %* | 47.3% | 54.5% | 0.1429 |
| Current smoker, % | 12.73% | 10.4% | 0.0736 |
| No. COVID-19 test (14 days), mean (SD) | 2.11 (1.7) | 2.18 (1.7) | 0.0421 |
| No. COVID-19 test (90 days), mean (SD) | 14.2 (8.9) | 14.6 (8.8) | 0.0398 |
| No. facility (+) tests (14 days), mean (SD) | 1.6 (3.4) | 1.4 (3.0) | 0.0679 |

\*COVID: coronavirus disease; DM: diabetes mellitus; MACE: major adverse cardiovascular events; ADRD: Alzheimers and related dementias; SMD= standardized mean difference

**Figure 4.**
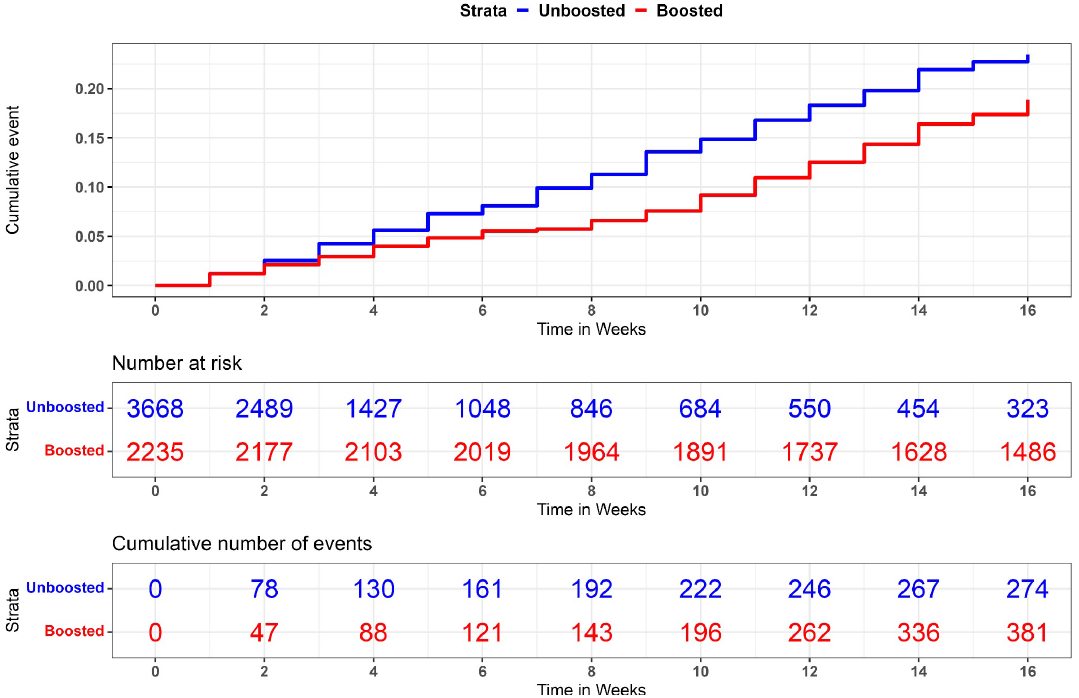
Km plots for infection following bivalent vaccination in comparison to controls in a TTE design.

**Figure 5.**
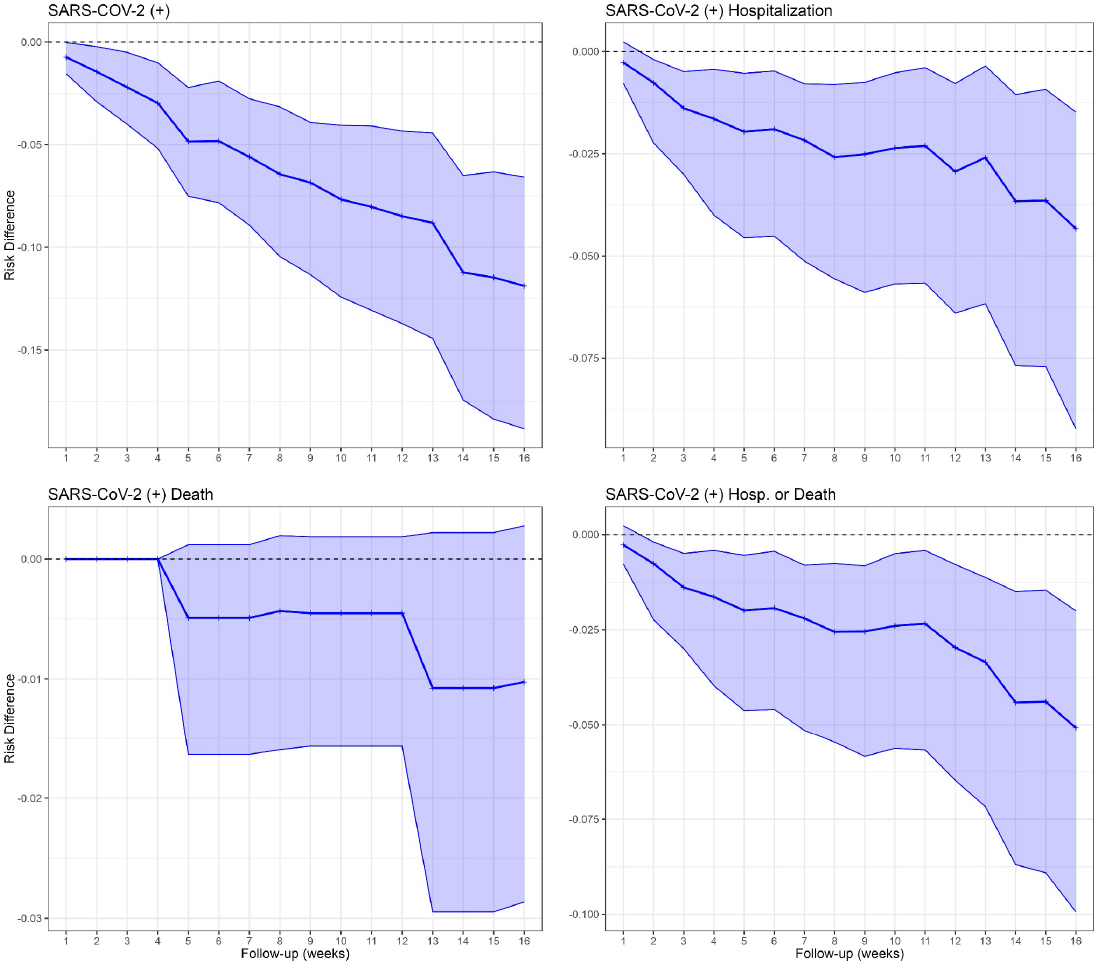
Risk differences for SARS-CoV-2 clinical outcomes of infection, hospitalization, death or a combination of hospitalization or death up to 16 weeks after bivalent vaccination.

**Table 5.** Unweighted and weighted relative risk reduction after bivalent vaccination over time.

| SARS-CoV-2 Outcome | 12 Week Unweighted | 12 Week Weighted | 16 Week Unweighted | 16 Week Weighted |
| --- | --- | --- | --- | --- |
| Laboratory-confirmed Infection | 31.5%<br>(18.4%, 42.7%) | 41.0%<br>(24.7%, 54.7%) | 19.5%<br>(5.8%, 32.1%) | 39.7%<br>(25.2%, 52.1%) |
| Hospitalization following infection | 35.5%<br>(12.1%, 53.7%) | 46.0%<br>(16.6%, 67.5%) | 26.9%<br>(3.3%, 44.4%) | 47.2%<br>(22.3%, 67.6%) |
| Death following infection | 56.8%<br>(-95.4%, 92.6%) | 70.2%<br>(-285.0%, 96.9%) | 52.8%<br>(-68.3%, 84.3%) | 74.4%<br>(-180.4%, 94.4%) |
| Hospitalization or death after infection | 37.1%<br>(14.3%, 55.8%) | 45.6%<br>(16.9% , 67.0%) | 30.7%<br>(8.8% , 47.1%) | 50.5%<br>(26.1%, 68.6%) |

\* With the exception of our follow-up time decisions these results mimic the methodology previously published (15).

## Discussion

This is among the first studies to evaluate immunity and clinical protection of the bivalent SARS-CoV-2 vaccination among nursing home residents. We show immunologic and clinical evidence of protection from two separate populations living in nursing homes. Immunologically, we show sustained if also waning evidence of elevated antibody and neutralizing activity six months after the bivalent vaccination. While the clinical data using a target trial emulation has an adequate sample size with up to four-months follow-up, it also supports that clinical benefit against severe outcomes persists many months after bivalent boosting. Together, our data demonstrate substantial evidence of protection through both approaches lasting four to six months after bivalent vaccination administration.

Neutralization assays provide a functional biologic correlate of protection, where higher titers indicate better protection. We show a continued rise in peak neutralization titers with each boost that wanes between boosts. Those without prior SARS-CoV-2 infection get a larger relative titer rise with each boost than those with prior infection, but the previously SARS-CoV-2 infected achieve even higher peak titers. Even from the prior two monovalent boosters, the BA.5 neutralization activity increases with each boost suggesting a further broadening of immunity to Omicron strains with each boost.

Because we observed no ceiling in the neutralization titers even after a third vaccination at least six months after the prior one, we would expect the addition of a bivalent vaccination similarly spaced will again produce substantially higher titers to at least match if not exceed the prior maximum neutralization activity.

Even though our SARS-CoV-2 naive nursing home resident population can maintain their BA.5 neutralization titers above the LLD for six months following their bivalent vaccination, their titers have fallen fivefold further compared to those with prior infection. Almost all individuals who had breakthrough infection had titers below 100 pNT50 (data not shown), indicating the greater vulnerability that occurs with the lowest titers. We have not formally determined an immune correlate for protection, and even high titers with or without prior infection will not prevent an infection from a large inoculum. These complicate the interpretation of neutralization titers in the guidance for a universal second bivalent booster recommendation.

The clinical TTE data shows protective benefit of bivalent vaccine to severe outcomes of hospitalization and death in the nursing home population. The subject follow-up time and events with this approach suffice for us to assess relative vaccine effectiveness up to 16 weeks following the bivalent vaccination. The short follow-up time represents a limitation of our study design and does not preclude a longer lasting benefit, even if the data suggest waning benefit after three months. We need to wait until more of the population has been followed for a longer period to fairly assess whether benefits continue to accrue or not; we will extend our analysis in future work. Several other studies have initial effectiveness data after the bivalent vaccination. Lin et al published in the general population (17) and Arbel et al in persons over age 65 (18) and both found significant benefit from BV vaccinations. Arbel et al’s study population was data from one of Israel’s large public health services and not focused on long term care residents. Our data is novel for looking at the focused population of long stay nursing home residents. The TTE has few women represented in this largely male Veteran population, limiting generalizability.

Here we endeavor to integrate the clinical and immunologic data to formulate considerations for when nursing home residents should receive their next booster. The strongest recommendation based on the clinical work and CDC (19) recommendation is that those who have never received a bivalent vaccination should consider receiving the bivalent vaccine as soon as possible. The immunology also strongly supports this recommendation between this study and our prior studies on NH residents (2, 3, 9-11). Waning immunity over time since those who received a bivalent vaccine shortly after approval may no longer have the expected clinical protection. By six months after the bivalent vaccination, while 80% of the group not previously infected with SARS-CoV-2 still had BA.5 neutralization titers above LLD, their titers were 1/5th of those who had previously recovered from infection. Subclinical SARS-CoV-2 infections represent the majority of breakthroughs in vaccinated individuals (20), introducing uncertainty into clinical ascertainment of who has had prior infection and therefore which nursing home residents have a stronger rationale to consider a booster. Natural infection and vaccinations individually and together have continued to raise the lowest titers over time. Thus, as immunity accrues, we would consequently expect that we can extend the length of time between vaccinations because of increasing durability of effective immune protection from clinically important infection. Yet, we will still need to confront new variants with immune evasive properties as they continue to arise. Also, SARS-CoV-2, unlike other beta coronaviruses and influenza, has circulated widely throughout warmer months outside of the typical respiratory viral season. This complicates any decision to recommend waiting for a seasonal SARS-CoV-2 vaccine update in the fall using the seasonal influenza model for vaccination.

Our clinical and immunologic data together suggest that at least a substantial segment of the nursing home population could derive benefit with a second bivalent vaccine offered after four months after their prior bivalent vaccination. Our data do not indicate that an additional vaccination risks lower peak neutralization activity, i.e., those who opt for a booster now and also again for a formulation designed for the fall offering should not experience a less effective response of the next vaccine. We do understand that assertively recommending a second bivalent booster now risks increasing reluctance to accept a fall vaccination that likely will offer an updated formulation. Currently, only 53% of NH residents have received the bivalent vaccine (21), and many did not receive both monovalent boosters previously offered. Consequently, those willing or even eager to receive a second bivalent vaccination now *and* who will likely accept a fall booster would be the best candidates for a second bivalent vaccination now. Nursing home leadership could also rapidly endorse and strengthen support for any bivalent booster recommendations if a new variant with a large rise in cases sweeps in nationally. Our experience with each of the prior boosts (3, 10, 11) indicate that the benefits of an additional vaccination begin to accrue quickly--within weeks--supporting this watchful waiting strategy for those who might be hesitant now.

In the end, we agree that the FDA and CDC have approved the option for individuals to receive a second bivalent vaccination now. This approach allows maximum flexibility to residents and their caregivers on how they weigh their risk for severe illness, exposure risk due to their behaviors and use of PPE, general viral circulation, and the need to protect against SARS-CoV-2 infection and its consequences.

## Data Availability

All data produced in the present study are available upon reasonable request to the authors after it is published in a peer-reviewed journal.

## Funding

This work was supported by NIH AI129709-03S1, CDC 200-2016-91773, U01 CA260539-01, VA BX005507-0 and VA HSRD CIN 13-419

The views and opinions expressed are those of the authors and do not represent the policy of the US Dept of Veterans Affairs.

## Acknowledgments

Thank-you for these other individuals for assisting in various parts of the study.

Case Western Reserve University: Debbie Keresztesy, Dennis Wilk, Alexandra Paxitzis, Vaishnavi Ragavapuram, Nicholas Sundheimer, Htin Aung

Brown University & Lifespan: Rosa Baier, Evan Dickerson, Laurel Holland, Shreya Kamojjala, Eleftherios Mylonakis, Aman Nanda, Clare Nugent, Igor Vishnepolskiy, Tiffany Wallace

